# Transitions in ENDS and cigarette use among youth in the PATH Study from 2015–2023: a multistate transition modeling analysis

**DOI:** 10.64898/2026.04.14.26349857

**Authors:** Olivia K. Roberts, Jihyoun Jeon, Evelyn Jimenez-Mendoza, Stephanie R. Land, Neal D. Freedman, Rossana Torres-Alvarez, Ritesh Mistry, David T. Levy, Rafael Meza, Andrew F. Brouwer

## Abstract

**Introduction:** Monitoring trends in transitions in the use of electronic nicotine delivery systems (ENDS) and cigarettes among youth is important for understanding the potential public health impacts of these products.

**Methods:** Using a weighted Markov multistate transition model accounting for complex survey design, we estimated transition rates and one-year transition probabilities between never, non-current, ENDS-only, and cigarette use (with or without dual use of ENDS) among 26,744 youth aged 12–17 years who participated in at least two consecutive waves from Waves 2–7.5 (approximately 2015–2023) of the nationally representative Population Assessment of Tobacco and Health (PATH) Study. We also estimated transitions stratified by ages 12–14 and 15–17 years.

**Results:** The one-year probability of ENDS-only initiation from never use among youth peaked in 2017–19 (Waves 4–5) at 4.0% (95%CI: 3.6–4.3%) and was higher for 15–17-year-olds at 5.8% (95%CI: 5.2–6.4%) than 12–14-year-olds at 2.2% (95%CI: 1.8–2.6%). In the following years, ENDS-only initiation rates declined and plateaued, with 2.6% (95%CI: 2.3–3.0%) initiation in 2022–23. Cigarette initiation from never use decreased over 2015– 23 from 0.8% (95%CI: 0.6–1.0%) in 2015–16 to 0.1% (95%CI: 0.0–0.2%) in 2022–23. There was an increase in the fraction of youth who transitioned from non-current product use to ENDS-only use from 13.7% (95%CI: 7.5–20.0%) in 2015–16 to 35.1% (95%CI: 25.4–44.8%) in 2022–23, paired with a decrease in non-current use to cigarette use from 20.9% (95%CI: 11.8–30.0%) to 6.3% (95%CI: 1.7–10.8%). Transitions from ENDS-only or cigarette use to non-current use remained relatively constant over time at around 25% and 15% per year, respectively.

**Conclusion:** ENDS-only use initiation has changed over time, peaking around 2019 and subsequently decreasing and plateauing, but cessation rates for both ENDS and cigarettes have remained relatively stable. Thus, interruption of tobacco product initiation may be the most effective approach to reducing tobacco product use among youth.

**What this paper adds:** What is already known on this topic:

- Transitions in cigarette and ENDS use have changed over time, with youth more likely to adopt ENDS and less likely to adopt cigarettes than older age groups.

What this study adds

- We found that ENDS initiation among youth peaked around 2019 and was higher for those 15–17 years than 12–14 years. There were few significant differences between the two age groups for other transitions.
- Cigarette initiation among youth declined over this period. Cessation rates for both ENDS and cigarettes have remained relatively stable.

How this study might affect research, practice or policy

- Tobacco control efforts should prioritize preventing all tobacco and nicotine product initiation among youth.

## Introduction

Sales of electronic cigarettes and other electronic nicotine delivery systems (ENDS) increased substantially beginning in 2017, with the increased prevalence of JUUL-brand use, particularly among youth.^1-3^ Although promoted as a harm reduction and cigarette cessation tool for adults who smoke, there remain concerns that the availability and design of ENDS products may appeal to youth, leading to nicotine addiction and possible cigarette initiation.^4-6^ Indeed, youth perceive many ENDS design aspects, including flavors, as targeted to them.^7^ Continued monitoring of transitions in ENDS use among youth is necessary for understanding the changing public health impact of ENDS, projecting future use, and developing interventions. Previously, we applied a multistate transition model to the nationally representative longitudinal Population Assessment of Tobacco and Health (PATH) Study to analyze how transitions among ENDS and cigarette use changed for youth between 2015–17 and 2017–19 (Waves 4–5).^8^ In this analysis, we analyzed youth product use transitions across the broader 2015–2023 (Waves 2–7.5) period. Our multistate transition modeling approach allows us to estimate the underlying transition rates that lead to tobacco and nicotine use patterns among youth and characterize patterns and trends in these transitions over time.

## Methods

### Data

We used data on 26,744 total youth from Waves 2–7.5 (2015–2023) of the PATH Study, including special collections Waves 4.5, 5.5, and 7.5. Wave 1 was not included in our main analysis because the ENDS established use criterion, discussed below, was not available for youth in Wave 1.

Follow-up time between Waves was assumed to be one or two years, as appropriate for each Wave pair. We categorized participants into age groups 12–14 and 15–17 years. If youth were 17 years old in a given Wave, we included their data at the follow-up Wave (where they would be age 18 or 19) but did not include them in subsequent Waves, ensuring we included transition estimates for 17-year-olds but not for young adults.

Each participant’s product use was categorized as never established use (of either product), non-current use, ENDS-only use, or cigarette use (with or without dual use of ENDS). Established ENDS use was defined as “fairly regular” use of ENDS, while established cigarette use was defined as 100+ lifetime cigarettes. Current ENDS-only use was defined as established and past 30-day use of ENDS, while current cigarette use was defined as established and past 30-day use of cigarettes, with or without past 30-day use of ENDS, following the approach in the Smoking and Vaping Model (SAVM).^9^ Dual use was not modeled as a separate category due of the low number of participants only smoking cigarettes in more recent Waves. Non-current use was defined as no past 30-day use of either product by a participant who previously had established use of one or both products. In this analysis, we do not refer to this category as “former” use, because former use has connotations of longer-term cessation, whereas our categories are defined by past-30-day use. See Figure S1 in the Supplementary Material for state definitions and allowed transitions between states. Characteristics of the population by Wave are given in Tables S1 and S2.

Our main analysis focused on the categories using the established use definitions, so that experimental use was not considered current use of that product. We also performed a sensitivity analysis in which the categories were defined by current use only, without regard to established use. Because this analysis did not use the established use questions, we applied it to Waves 1–7.5 (2014–2023). The characteristics of the population for this analysis can be found in the Supplementary Material (Tables S3 and S4).

### Transition modeling

We used a Markov multistate transition model to analyze the underlying transition hazard rates between product use for youth overall and in each of the two age categories (12–14 and 15–17 years). Multistate transition models assume transitions arise from a continuous-time, finite-state stochastic process where the transition rates depend only on the current state, not on past states or transition history. Technical details are provided in the Supplementary Material. The model estimated transition hazard rates, which are the instantaneous risks of transitioning from one state to another. We incorporated longitudinal survey weights for each Wave transition into the model. In addition to the rates estimates for all adjacent Wave pairs, we also estimated transition rates for Wave pairs 4–5 and 5–6 because the special collection Waves between them excluded some ages: Wave 4.5 excluded 18–year-olds, and Wave 5.5 excluded 12–year-olds, so the 15–17 and 12–14 age groups were excluded for the Waves 4–4.5 (2017-18) and Waves 5.5–6 (2020-21) transitions, respectively. Multistate transition models were estimated using the wmsm function^10^ in R (available at https://tcors.umich.edu/Resources_Research.php), which is an extension of the msm function^11^ modified to incorporate participant weights. This analysis of publicly available data was not regulated as human subjects research (University of Michigan Institutional Review Board HUM00162265).

Using the estimated transition rates, we estimated the one-year transition probabilities for each group for each pair of Waves separately. For Wave pairs two years apart, the one-year transition probabilities represent estimated transitions per year over the two-year period. We censored any transition estimate that was informed by fewer than 5 individuals because of high uncertainty.

## Results

There were statistically significant changes in the one-year transition probabilities over time (Figure 1). Among youth who never used either product, ENDS-only use initiation peaked between 2017–19 (Waves 4–5) at 4.0% (95%CI: 3.6–4.3%) and was higher for 15–17-year-olds at 5.8% (95%CI: 5.2–6.4%) than 12–14-year-olds at 2.2% (95%CI: 1.8–2.6%) (Figure 2). In the following years, ENDS initiation declined and plateaued, with initiation of 2.6% (95%CI: 2.3– 3.0%) overall in 2022–23 (Waves 7–7.5). Among those who used ENDS only, the transition to non-current use remained relatively constant after 2017, at around 25% per year, with no difference by age group.

**Figure 1.**
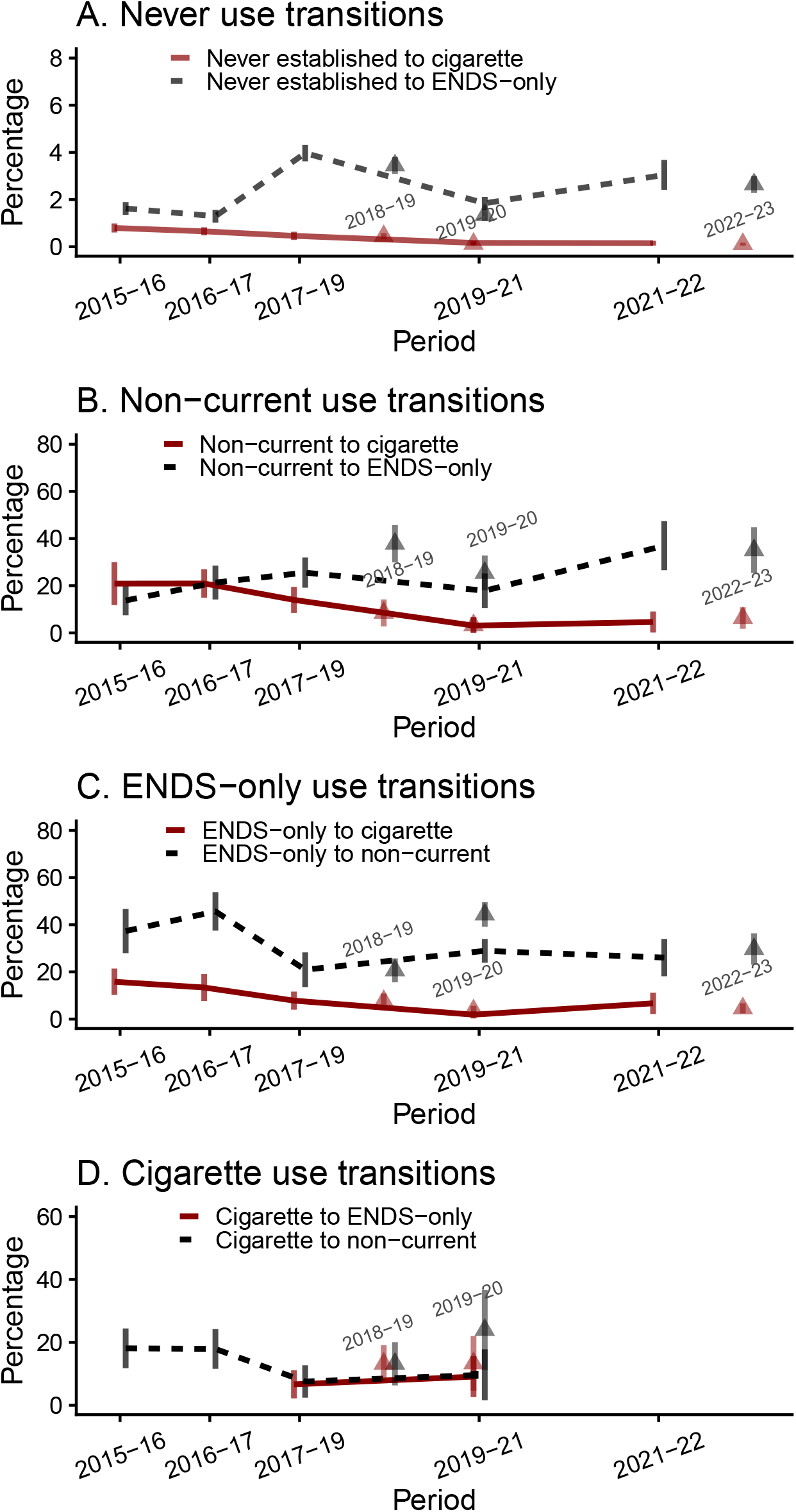
One-year transition probabilities and 95% confidence intervals for youth, PATH Waves 2–7.5 (2015–23), by product use state. The x-axis represents the wave pair, and the y-axis represents the modeled probability of transition in one year. Connected points indicate standard transitions between main Waves, while triangular points correspond to transitions involving special collection Waves: 4.5–5 (2018–19), 5–5.5 (2019–20), and 7–7.5 (2022–23). Results based on fewer than five observed transitions were censored because of high uncertainty.

**Figure 2.**
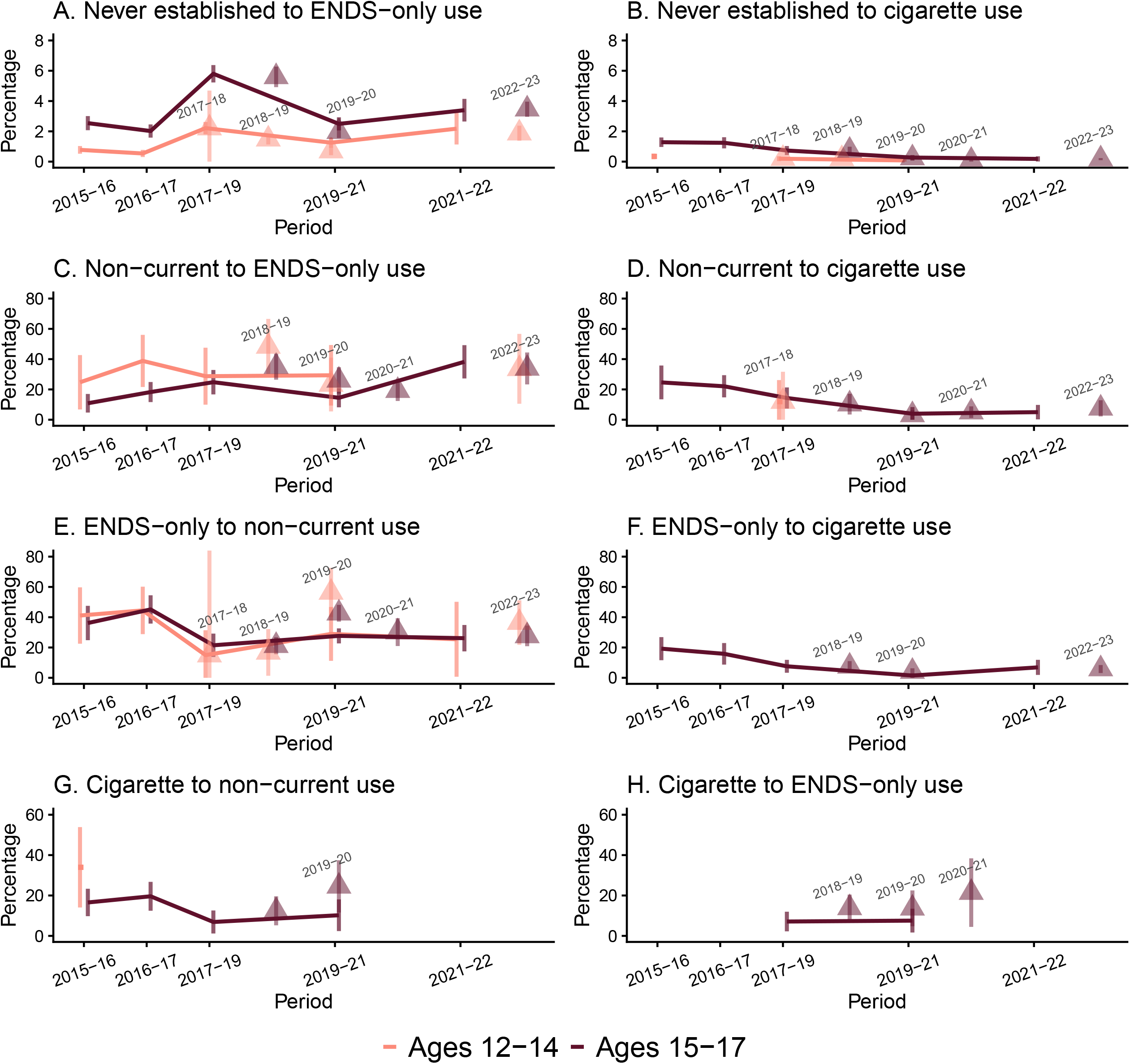
One-year transition probabilities and 95% confidence intervals for youth, PATH Waves 2–7.5 (2015–23), by transition and age group. The x-axis represents the year of the starting wave in each transition, and the y-axis represents the modeled probability of transition in one year. Connected and square points indicate standard transitions between main Waves, while triangular points correspond to transitions involving special collection Waves: 4–4.5 (2017–18), 4.5–5 (2018–19), 5–5.5 (2019–20), 5.5–6 (2020–21), and 7–7.5 (2022–23). Results based on fewer than five observed transitions were censored because of high uncertainty.

The one-year probability of cigarette initiation from never use of either product decreased over 2015–23 from 0.8% (95%CI: 0.6–1.0%) in 2015–16 to 0.1% (95%CI: 0.0–0.2%) in 2022–23. Transitions from cigarette use to non-current use of either product remained relatively steady at about 15% per year, regardless of age group. Among non-current users, relapse to ENDS-only and cigarette use occurred at much higher rates than primary initiation. The percentage of youth who transitioned to ENDS-only use from non-current use in one year increased overall over the period from 13.7% (95%CI: 7.5–20.0%) in 2015–16 to 35.1% (95%CI: 25.4–44.8%) in 2022–23, paired with a decreasing trend in transitions to cigarette use from non-current use from 20.9% (95%CI: 11.8–30.0%) to 6.3% (95%CI: 1.7–10.8%).

Although 15–17-year-olds consistently initiated ENDS-only use at higher rates than 12–14-year-olds in every pair of Waves, no other transition showed consistent differences between the two age groups. Even though the older age group initiated ENDS at higher rates, both age groups initiated cigarettes (with or without ENDS use) at similar rates and cessation rates were similar for both age groups. Transitions for the 12–14-year-old age group are more frequently censored because of lower prevalence of established product use.

The results of our sensitivity analysis, which included experimental cigarette and ENDS use (i.e., removing the established use criteria for cigarettes and ENDS from our product use state definitions), are presented in Figures S1 and S2 in the Supplementary Material. Because the prevalence of current product use is higher when including experimental use, there is less censoring of transitions, but the transition rates are generally comparable to our main analysis.

## Discussion

We applied a weighted multistate transition model to estimate transition rates for the initiation and cessation of different tobacco products among youth in the PATH study from 2015 to 2023. We found that ENDS-only use initiation peaked around 2019 and subsequently plateaued, while cigarette initiation (with or without ENDS use) has decreased over the period. Relapse rates of either or both products among those not currently using either product remained higher than primary initiation rates but increased over time for ENDS-only use and decreased for cigarette use. Transitions from either or both products to non-current use have remained relatively stable over time. However, the probability of transitioning from ENDS-only to non-current use was consistently higher than transition rates from cigarette use to non-current use, illustrating the more transient nature of ENDS-only use among youth.

There have been several shifts in the youth tobacco use landscape during this period that may have influenced these patterns. In 2018, the U.S. Surgeon General declared an ENDS use epidemic among youth and advised immediate action among public health professionals.^12^ Increased awareness of the health risks associated with vaping, especially during the 2019–20 lung injury outbreak, likely shaped youth perceptions, as studies show that youth who heard about the lung injury outbreak were more likely to consider vaping harmful.^13-14^ Following these events, policymakers heightened regulatory strategies, including e-cigarette flavor bans, tobacco 21 laws, and anti-vaping campaigns.^15-17^ These efforts may have contributed to changes in initiation trends.

Multistate transition modeling is increasingly used in tobacco control to estimate transition rates between states in longitudinal studies.^18-19^ However, a limitation of multistate transition models is the assumption that rates depend on the current state, not past states. This prevents an individual’s full use history from being considered. For example, an individual who has been using tobacco products for a longer period may be less likely to successfully quit using those products, but the model does not take this into account. The model assumes the transition probabilities are the same regardless of past states or duration of past states. Our results should be viewed as population averages. Additionally, because of the small number of observations for some transitions, some transition estimates were censored, particularly for transitions to and from cigarette use, reflecting the low prevalence of established cigarette use among youth in more recent Waves. A further limitation of the model is that the use of other tobacco or nicotine products is not included. In particular, nicotine pouch use and dual use with e-cigarettes among youth increased from 2023 to 2024.^20^ Nicotine pouch use may have had an impact on some of the most recent Wave transitions in our analysis.

Low rates of cigarette initiation are a noteworthy finding from our study. As an observational study, however, we are unable to determine whether such low rates reflect the introduction of ENDS or other tobacco or nicotine products into the market, long-term trends in cigarette use, or other factors. There is some evidence that frequent ENDS use among youth may be a catalyst for future cigarette use, and that youth are not likely to use ENDS products to aid cigarette cessation.^21^ Conversely, ecological evidence has found decreasing cigarette prevalence even as e-cigarette prevalence has climbed among youth.^22^ Indeed, our analysis suggests that cigarette initiation rates have dropped while 30-day cessation rates have remained stable despite rising ENDS use, and ENDS to cigarette use transitions have remained low.

Our results highlight the importance of tobacco control efforts aimed at reducing initiation of all tobacco products among youth. Although cessation rates have not improved, a decline in cigarette initiation and a plateau in ENDS initiation suggest that the changing marketplace and public health interventions may be shifting youth product use patterns. Continued monitoring of transitions could help inform appropriate public health interventions for youth as the product landscape continues to evolve.

## Data Availability

Public Use Files from the Population Assessment of Tobacco and Health Study are available for download from an open access repository (https://doi.org/10.3886/ICPSR36498.v23).

## Acknowledgments

This project was funded through National Cancer Institute (NCI) and Food and Drug Administration (FDA) grant U54CA229974. The opinions expressed in this article are the authors’ own and do not necessarily reflect the views of the National Institutes of Health, the Department of Health and Human Services, or the United States government.

## Competing Interests

All authors declare that they have no competing interests.

## Contributors

Conceptualization and methodology: AFB and RMe; data curation: JJ and EJ-M; analysis and original draft preparation: OKR; review and editing: JJ, EJ-M, SRL, NDF, RTA, RMi, DTL, RMe, and AFB; funding acquisition: RMe and DTL; guarantor: AFB.

## Data availability statement

Public Use Files from the Population Assessment of Tobacco and Health Study are available for download from an open access repository (https://doi.org/10.3886/ICPSR36498.v23). Conditions of use are available on the aforementioned websites.

## Notes

### Competing Interest Statement

The authors have declared no competing interest.

### Author Declarations

This study only used data that were publicly available. Public Use Files from the Population Assessment of Tobacco and Health Study are available for download from an open access repository (https://doi.org/10.3886/ICPSR36498.v23). This analysis of publicly available data was not regulated as human subjects research (University of Michigan Institutional Review Board HUM00162265).

