## Supplementary material for "Transitions in ENDS and cigarette use among youth in the PATH Study from 2015–2023: a multistate transition modeling analysis"

This technical appendix is reproduced in part from the supplementary material of Brouwer et al [10] but includes updated notation and technical details. Additional details may be found in Jackson [11], and a tutorial for working with the weighted multistate transition model (wmsm) code is available at <https://tcors.umich.edu/Resources_Research.php> along with the code itself.

A multistate transition model is a continuous-time, finite-state stochastic process with the Markov assumption that transition rates depend only on the current state and not on past states or transition history. We denote the state of the process at time $t$ as $S\left( t \right)$. We denote the probability that an individual is in state $j$ after an amount of $\Delta t$ since they were observed in state $i$ as

$$\begin{aligned} P_{ij}(t,t+\Delta t)=Pr\left[ S\left( t+\Delta t \right)=j | S\left( t \right)=i \right]. \#\left( 1 \right) \end{aligned}$$

In general, the transition probabilities can depend on the observation time $t$ in addition to the time span $\Delta t$, and in this analysis we consider discrete time periods over which we assume that that the model is homogeneous in time, i.e., $P_{ij}\left( t,t+\Delta t \right)= P_{ij}(0,\Delta t)$. Thus, we drop the dependence on $t$ moving forward and write $P_{ij}(\Delta t)$. We then define the hazard rate of the transition from state $i$ to state$j$, for $i\neq j$, as

$$\begin{aligned} q_{ij}=\lim_{\Delta t\to0} \frac{1}{\Delta t}Pr\left[ S\left( \Delta t \right)=j | S\left( 0 \right)=i \right]. \#\left( 2 \right) \end{aligned}$$

The transition hazard rates form a matrix $Q = \left[ q_{ij} \right],$ where the diagonal entries are given by $q_{ii}=-\sum_{j\neq i} q_{ij}$. The transition probability matrix $P\left( \Delta t \right)=\left[ P_{ij}\left( \Delta t \right) \right]$ is a function of the transition hazard rates and may be calculated as the matrix exponential of $\Delta t\cdot Q$, that is

$\begin{aligned} P\left( \Delta t \right)=\text{expm}\left( \Delta t\cdot Q \right).\#\left( 3 \right) \end{aligned}$

Transition probabilities can be estimated for any value of $\Delta t$, assuming that the transition hazards $q_{ij}$ do not change over that time period (assumption of homogeneity).

The values of the transition hazard rates $q_{ij}$of multistate transition model are estimated by maximizing a statistical likelihood $L$ given a set of observed states and times by comparing the observed states to the probabilities in $P\left( \Delta t \right)$ as a function of the transition hazard matrix $Q$. Specifically, consider a set of individuals $m = 1, \ldots,N$ and their observed states $s_{m,t_{m,k}}$at times $t_{m,k}$, where $k$ is the index of individual $m$’s $K_{m}$observations in the data. Denote the data as $s=\{s_{m,t_{m,k}}\}$. We assume the individuals are independent, and thus we multiply all the modeled probabilities of the observed transitions:

$$\begin{aligned} L\left( Q|s \right)=\prod_{m=1}^{N} \prod_{k=1}^{K_{m}-1} P_{s_{m,t_{m,k}},s_{m,t_{m,k+1}}}(t_{m,k+1}-t_{m,k}).\#\left( 4 \right) \end{aligned}$$

where $P_{i,j}$ is a function of $Q$ as in Eqn (3).

Participant weights $W_{m}$can be incorporated into a weighted likelihood $L^{*}$. Although it is not strictly necessary to normalize the weights to the population size, $w_{m}=N\cdot W_{m}/\sum_{v} W_{v}$, it is convenient to do so because the resulting likelihood will correspond to the unweighted likelihood when all participants have equal weight. The weighted likelihood is given by

$$\begin{aligned} L^{*}\left( Q|s \right)=\prod_{m=1}^{N} \prod_{k=1}^{K_{m}-1} \left( P_{s_{m,t_{m,k}},s_{m,t_{m,k+1}}}(t_{m,k+1}-t_{m,k}) \right)^{w_{m}}.\#\left( 5 \right) \end{aligned}$$

Following the msm package [32], we assume that estimated transition hazard rates $\hat{q}_{ij}$ are normally distributed on the log-scale, that is

$$\begin{aligned} \log\hat{q}_{ij}\sim N\left( \mu=\text{mean}\left( \log\hat{q}_{ij} \right),\sigma^{2}=V\left( \log\hat{q}_{ij} \right) \right),\#\left( 6 \right) \end{aligned}$$

where $V$ denotes the estimated variance.

We estimated weighted point estimates for the log transition hazard rates $\log\hat{q}_{ij}$ by minimizing ${-logL}^{*}\left( Q|s \right)$as a function of the transition hazard rates. (This approach is equivalent to maximum likelihood estimation).

Variance estimates $V\left( \log\hat{q}_{ij} \right)$are calculated using replicate weights $w_{m}^{r}$. Replicate weights are a way to account for complex survey design aspects, such as strata and primary sampling units. PATH uses a variant of balanced repeated replication called Fay’s method to calculate 100 replicate weights. We calculate $\log\hat{q}_{ij}^{r}$ for each $r$ as above. Then, we calculate the variance of $\log\hat{q}_{ij}^{r}$ as

$$\begin{aligned} V\left( \log\hat{q}_{ij} \right)=c\sum_{r=1}^{100} \left( \log\hat{q}_{ij}^{r}-\log\hat{q}_{ij} \right)^{2}\#\left( 7 \right) \end{aligned}$$

where $c=1/(100\left( 1-0.3 \right)^{2})$ as specified by PATH. See the table below for the replicate weight variables used for each transition in this analysis.

| Transition | Weight Variable |
| --- | --- |
| Waves 1-2 | R02_YA_PWGT |
| Waves 2-3 | R03_YA_AWGT |
| Waves 3-4 | R04_YA_A01WGT |
| Waves 4-4.5 | X04_YA_S04WGT |
| Waves 4.5-5 | R05_YA_S04WGT |
| Waves 4-5 | R05_YA_S04WGT |
| Waves 5-5.5 | X05_YA_A04WGT |
| Waves 5.5-6 | R06_YA_A04WGT |
| Waves 5-6 | R06_YA_A04WGT |
| Waves 6-7 | R07_YA_A04WGT |
| Waves 7-7.5 | X07_YA_S07WGT |

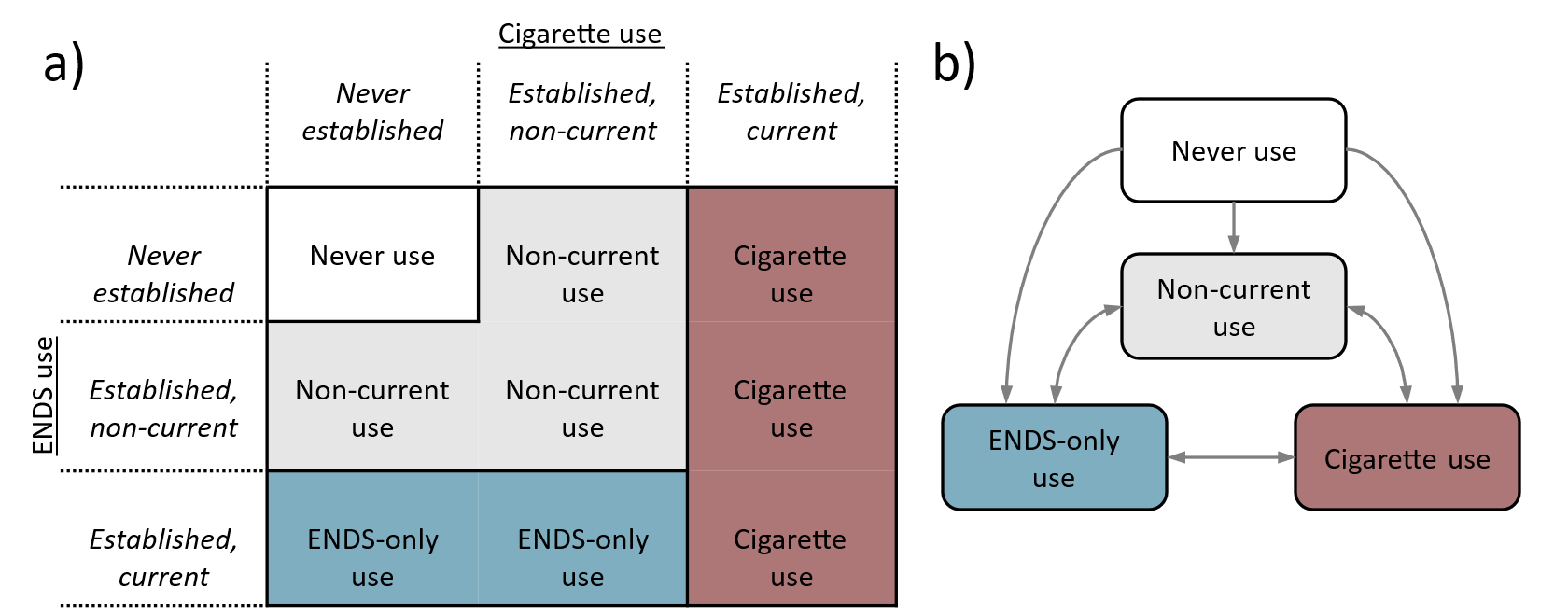

**Figure S1.** *a) Tobacco use state definitions. b) Direct transitions allowed between states in the model.*

***Table S1****: Characteristics of youth in the Population Assessment of Tobacco and Health (PATH) study with established use of cigarettes and ENDS in 2015–16 (Waves 2–3), 2016–17 (Waves 3–4), 2017–19 (Waves 4–5), 2019–21 (Waves 5–6), and 2021–22 (Waves 6–7) given as weighted percentages (%) and numbers (N). Note that in this table, “Tobacco & ENDS use state” is the prevalence for the earlier of the two waves.*

|  | Youth Waves 2-3 | | Youth Waves 3-4 | | Youth Waves 4-5 | | Youth Waves 5-6 | | Youth Waves 6-7 | |
| --- | --- | --- | --- | --- | --- | --- | --- | --- | --- | --- |
|  | % | N | % | N | % | N | % | N | % | N |
| Total | 100 | 11,098 | 100 | 10,487 | 100 | 12,764 | 100 | 9,018 | 100 | 4,582 |
| Gender |  |  |  |  |  |  |  |  |  |  |
| Female | 51.3 | 5,677 | 51.4 | 5,421 | 51.0 | 6,600 | 51.0 | 4,631 | 51.3 | 2,402 |
| Male | 48.7 | 5,421 | 48.6 | 5.066 | 49.0 | 6,164 | 49.0 | 4,387 | 48.7 | 2,180 |
| Race/ethnicity |  |  |  |  |  |  |  |  |  |  |
| Non-Hispanic White | 52.6 | 5,153 | 51.4 | 4,768 | 51.0 | 5,611 | 48.9 | 3,966 | 47.8 | 2,025 |
| Non-Hispanic Black | 22.5 | 3,204 | 23.1 | 3,044 | 23.0 | 3.809 | 23.9 | 2,623 | 24.4 | 1,322 |
| Hispanic | 13.1 | 1,447 | 12.6 | 1,363 | 12.8 | 1,676 | 12.1 | 1,101 | 11.9 | 516 |
| Non-Hispanic Other/Unknown | 11.8 | 1,294 | 12.9 | 1,312 | 13.2 | 1,668 | 15.1 | 1,328 | 15.9 | 719 |
| Age (years) |  |  |  |  |  |  |  |  |  |  |
| 12-14 | 50.7 | 5,713 | 50.4 | 5,348 | 49.7 | 6,426 | 50.8 | 3,994 | 27.5 | 1,042 |
| 15-17 | 49.3 | 5,385 | 49.6 | 5,139 | 50.3 | 6,338 | 49.2 | 5,024 | 72.5 | 3,540 |
| Tobacco & ENDS use |  |  |  |  |  |  |  |  |  |  |
| Never use | 96.1 | 10,684 | 95.7 | 10,066 | 96.3 | 12,294 | 94.1 | 8,463 | 94.4 | 4,325 |
| Non-current use | 1.2 | 137 | 1.5 | 155 | 1.4 | 183 | 1.4 | 132 | 2.2 | 104 |
| Cigarette-only use | 1.2 | 131 | 0.9 | 91 | 0.7 | 92 | 0.3 | 30 | 0.1 | 7 |
| Non-daily | 0.6 | 69 | 0.6 | 56 | 0.4 | 49 | 0.2 | 15 | 0.1 | 4 |
| Daily | 0.6 | 62 | 0.4 | 35 | 0.3 | 43 | 0.2 | 15 | 0.1 | 3 |
| ENDS-only use | 1.1 | 106 | 1.4 | 140 | 1.3 | 167 | 3.7 | 349 | 3.0 | 137 |
| Non-daily | 0.9 | 84 | 1.3 | 124 | 1.1 | 144 | 2.9 | 274 | 1.8 | 85 |
| Daily | 0.2 | 22 | 0.2 | 16 | 0.2 | 23 | 0.8 | 75 | 1.2 | 52 |
| Dual cigarette/ENDS user | 0.4 | 40 | 0.4 | 35 | 0.2 | 28 | 0.5 | 44 | 0.2 | 9 |
| Non-daily cigarette, non-daily ENDS | 0.2 | 19 | 0.2 | 17 | 0.1 | 16 | 0.2 | 15 | 0.1 | 2 |
| Non-daily cigarette, daily ENDS | <0.1 | 7 | <0.1 | 2 | <0.1 | 3 | 0.2 | 16 | 0.1 | 6 |
| Daily cigarette, non-daily ENDS | 0.1 | 10 | 0.1 | 12 | <0.1 | 5 | 0.1 | 8 | <0.1 | 1 |
| Daily cigarette, daily ENDS | <0.1 | 4 | <0.1 | 4 | <0.1 | 4 | <0.1 | 5 | 0 | 0 |

***Table S2****: Characteristics of youth in the Population Assessment of Tobacco and Health (PATH) study with established use of cigarettes and ENDS in 2017–18 (Waves 4–4.5), 2018–19 (Waves 4.5–5), 2019–20 (Waves 5–5.5), 2020–21 (Waves 5.5–6), and 2022–23 (Waves 7–7.5) given as weighted percentages (%) and numbers (N). Note that in this table, “Tobacco & ENDS use state” is the prevalence for the earlier of the two waves.*

|  | Youth Waves 4-4.5 | | Youth Waves 4.5-5 | | Youth Waves 5-5.5 | | Youth Waves 5.5-6 | | Youth Waves 7-7.5 | |
| --- | --- | --- | --- | --- | --- | --- | --- | --- | --- | --- |
|  | % | N | % | N | % | N | % | N | % | N |
| Total | 100 | 6,647 | 100 | 11,871 | 100 | 9,221 | 100 | 4,087 | 100 | 8,562 |
| Gender |  |  |  |  |  |  |  |  |  |  |
| Female | 51.2 | 3,498 | 51.0 | 6,163 | 51.1 | 4,746 | 50.7 | 2,116 | 51.4 | 4,476 |
| Male | 48.8 | 3,149 | 49.0 | 5,708 | 48.9 | 4,475 | 49.3 | 1,971 | 48.6 | 4,086 |
| Race/ethnicity |  |  |  |  |  |  |  |  |  |  |
| Non-Hispanic White | 49.6 | 2,920 | 50.5 | 5,308 | 49.1 | 4,135 | 48.8 | 1,775 | 47.2 | 3,970 |
| Non-Hispanic Black | 23.3 | 1,933 | 23.1 | 3,462 | 23.8 | 2,622 | 23.5 | 1,187 | 24.9 | 2,490 |
| Hispanic | 12.4 | 815 | 12.2 | 1,476 | 12.2 | 1,115 | 11.7 | 481 | 12.4 | 940 |
| Non-Hispanic Other/Unknown | 14.8 | 979 | 14.2 | 1,625 | 14.9 | 1,349 | 15.9 | 644 | 15.5 | 1,162 |
| Age (years) |  |  |  |  |  |  |  |  |  |  |
| 12-14 | 100 | 6,647 | 50.3 | 5,609 | 50.5 | 4,061 | 0 | 0 | 50.9 | 3,387 |
| 15-17 | 0 | 0 | 49.7 | 6,262 | 49.5 | 5,160 | 100 | 4,087 | 49.1 | 4,725 |
| Tobacco & ENDS use |  |  |  |  |  |  |  |  |  |  |
| Never use | 99.1 | 6,582 | 95.2 | 11,288 | 93.8 | 8,636 | 92.7 | 3,798 | 95.5 | 8,139 |
| Non-current use | 0.5 | 33 | 1.2 | 148 | 1.5 | 143 | 3.0 | 128 | 1.3 | 125 |
| Cigarette-only use | 0.1 | 7 | 0.6 | 79 | 0.3 | 30 | 0.2 | 11 | <0.1 | 5 |
| Non-daily | 0.1 | 5 | 0.4 | 54 | 0.1 | 15 | 0.1 | 4 | <0.1 | 4 |
| Daily | <0.1 | 2 | 0.2 | 25 | 0.2 | 15 | 0.2 | 7 | <0.1 | 1 |
| ENDS-only use | 0.3 | 22 | 2.7 | 319 | 4.1 | 376 | 3.8 | 141 | 2.9 | 276 |
| Non-daily | 0.2 | 20 | 2.2 | 261 | 3.2 | 295 | 2.6 | 94 | 2.1 | 191 |
| Daily | <0.1 | 2 | 0.5 | 58 | 0.9 | 81 | 1.2 | 47 | 0.9 | 85 |
| Dual cigarette/ENDS user | <0.1 | 3 | 0.3 | 37 | 0.4 | 36 | 0.2 | 9 | 0.2 | 17 |
| Non-daily cigarette, non-daily ENDS | <0.1 | 3 | 0.2 | 20 | 0.1 | 15 | 0.1 | 3 | 0.1 | 8 |
| Non-daily cigarette, daily ENDS | 0 | 0 | 0.1 | 8 | 0.1 | 12 | 0.1 | 3 | 0.1 | 7 |
| Daily cigarette, non-daily ENDS | 0 | 0 | 0.1 | 8 | <0.1 | 5 | 0.1 | 2 | <0.1 | 1 |
| Daily cigarette, daily ENDS | 0 | 0 | <0.1 | 1 | <0.1 | 4 | <0.1 | 1 | <0.1 | 1 |

***Table S3****: Characteristics of youth in the Population Assessment of Tobacco and Health (PATH) study with experimental use of cigarettes and ENDS in 2014–15 (Waves 1–2), 2015–16 (Waves 2–3), 2016–17 (Waves 3–4), 2017–19 (Waves 4–5), 2019–21 (Waves 5–6), and 2021–22 (Waves 6–7) given as weighted percentages (%) and numbers (N). Note that in this table, “Tobacco & ENDS use state” is the prevalence for the earlier of the two waves.*

|  | Youth Waves 1-2 | | Youth Waves 2-3 | | Youth Waves 3-4 | | Youth Waves 4-5 | | Youth Waves 5-6 | | | Youth Waves 6-7 | |
| --- | --- | --- | --- | --- | --- | --- | --- | --- | --- | --- | --- | --- | --- |
|  | % | N | % | N | % | N | % | N | % | N | | % | N |
| Total | 100 | 11,749 | 100 | 11,073 | 100 | 10,450 | 100 | 12,692 | 100 | | 8,961 | 100 | 4,539 |
| Gender |  |  |  |  |  |  |  |  |  | |  |  |  |
| Female | 51.1 | 5,993 | 51.3 | 5,664 | 51.4 | 5,408 | 51.0 | 6,564 | 51.0 | | 4,602 | 51.2 | 2,381 |
| Male | 48.9 | 5,756 | 48.7 | 5,409 | 48.6 | 5,042 | 49.0 | 6,128 | 49.0 | | 4,359 | 48.8 | 2,158 |
| Race/ethnicity |  |  |  |  |  |  |  |  |  | |  |  |  |
| Non-Hispanic White | 53.8 | 5,598 | 52.7 | 5,147 | 51.4 | 4,748 | 51.1 | 5,585 | 49.0 | | 3,948 | 47.9 | 2,015 |
| Non-Hispanic Black | 22.0 | 3,340 | 22.5 | 3,197 | 23.1 | 3,036 | 22.9 | 3,779 | 23.9 | | 2,611 | 24.5 | 1,314 |
| Hispanic | 13.4 | 1,541 | 13.0 | 1,441 | 12.6 | 1,362 | 12.8 | 1,673 | 12.1 | | 1,094 | 11.9 | 506 |
| Non-Hispanic Other/Unknown | 10.8 | 1,270 | 11.8 | 1,288 | 12.9 | 1,304 | 13.2 | 1,655 | 15.0 | | 1,308 | 15.7 | 704 |
| Age (years) |  |  |  |  |  |  |  |  |  | |  |  |  |
| 12-14 | 50.4 | 6,079 | 50.6 | 5,692 | 50.4 | 5,328 | 49.7 | 6,388 | 50.7 | | 3,964 | 27.5 | 1,035 |
| 15-17 | 49.6 | 5,670 | 49.4 | 5,381 | 49.6 | 5,122 | 50.3 | 6,304 | 49.3 | | 4,997 | 72.5 | 3,504 |
| Tobacco & ENDS use |  |  |  |  |  |  |  |  |  | |  |  |  |
| Never use | 83.5 | 9,830 | 79.8 | 8,860 | 81.0 | 8,505 | 82.2 | 10,455 | 80.0 | | 7,076 | 81.6 | 3,697 |
| Non-current use | 10.6 | 1,244 | 14.4 | 1,602 | 13.5 | 1,406 | 11.9 | 1,523 | 11.3 | | 1,070 | 12.6 | 585 |
| Cigarette-only use | 3.1 | 356 | 2.5 | 268 | 1.8 | 174 | 1.8 | 240 | 0.8 | | 75 | 0.3 | 15 |
| Non-daily | 2.5 | 283 | 1.9 | 211 | 1.5 | 142 | 1.6 | 203 | 0.6 | | 62 | 0.2 | 11 |
| Daily | 0.6 | 73 | 0.5 | 57 | 0.3 | 32 | 0.3 | 37 | 0.1 | | 13 | 0.1 | 4 |
| ENDS-only use | 1.5 | 169 | 2.0 | 203 | 2.6 | 250 | 3.0 | 346 | 6.5 | | 611 | 4.8 | 211 |
| Non-daily | 1.5 | 164 | 1.9 | 189 | 2.5 | 238 | 2.8 | 328 | 5.8 | | 545 | 3.7 | 165 |
| Daily | 0.1 | 5 | 0.1 | 14 | 0.1 | 12 | 0.2 | 18 | 0.7 | | 66 | 1.1 | 46 |
| Dual cigarette/ENDS user | 1.2 | 150 | 1.3 | 140 | 1.2 | 115 | 1.1 | 128 | 1.4 | | 129 | 0.7 | 31 |
| Non-daily cigarette, non-daily ENDS | 1.0 | 114 | 1.0 | 99 | 0.8 | 81 | 0.8 | 96 | 0.8 | | 80 | 0.4 | 16 |
| Non-daily cigarette, daily ENDS | 0.1 | 9 | 0.1 | 16 | 0.1 | 7 | 0.1 | 12 | 0.3 | | 30 | 0.3 | 14 |
| Daily cigarette, non-daily ENDS | 0.2 | 26 | 0.2 | 21 | 0.3 | 23 | 0.1 | 16 | 0.1 | | 14 | <0.1 | 1 |
| Daily cigarette, daily ENDS | <0.1 | 1 | <0.1 | 4 | <0.1 | 4 | <0.1 | 4 | 0.1 | | 5 | 0 | 0 |

***Table S4****: Characteristics of youth in the Population Assessment of Tobacco and Health (PATH) study with experimental use of cigarettes and ENDS in 2017–18 (Waves 4–4.5), 2018–19 (Waves 4.5–5), 2019-20 (Waves 5–5.5), 2020–21 (Waves 5.5–6), and 2022–23 (Waves 7–7.5) given as weighted percentages (%) and numbers (N). Note that in this table, “Tobacco & ENDS use state” is the prevalence for the earlier of the two waves.*

|  | Youth Waves 4-4.5 | | Youth Waves 4.5-5 | | Youth Waves 5-5.5 | | Youth Waves 5.5-6 | | Youth Waves 7-7.5 | | |
| --- | --- | --- | --- | --- | --- | --- | --- | --- | --- | --- | --- |
|  | % | N | % | N | % | N | % | N | % | N | |
| Total | 100 | 6,622 | 100 | 11,808 | 100 | 9,153 | 100 | 4,043 | 100 | | 8,497 |
| Gender |  |  |  |  |  |  |  |  |  | |  |
| Female | 51.1 | 3,485 | 51.0 | 6,127 | 51.1 | 4,708 | 50.5 | 2,088 | 51.3 | | 4,435 |
| Male | 48.9 | 3,137 | 49.0 | 5,681 | 48.9 | 4,445 | 49.5 | 1,955 | 48.7 | | 4,062 |
| Race/ethnicity |  |  |  |  |  |  |  |  |  | |  |
| Non-Hispanic White | 49.6 | 2,913 | 50.6 | 5,287 | 49.3 | 4,117 | 49.0 | 1,765 | 47.2 | | 3,943 |
| Non-Hispanic Black | 23.2 | 1,920 | 23.1 | 3,438 | 23.8 | 2,607 | 23.6 | 1,176 | 24.9 | | 2,469 |
| Hispanic | 12.4 | 813 | 12.2 | 1,471 | 12.1 | 1,104 | 11.6 | 472 | 12.4 | | 932 |
| Non-Hispanic Other/Unknown | 14.8 | 976 | 14.1 | 1,612 | 14.8 | 1,325 | 15.8 | 630 | 15.5 | | 1,153 |
| Age (years) |  |  |  |  |  |  |  |  |  | |  |
| 12-14 | 100 | 6,622 | 50.4 | 5,588 | 50.6 | 4,035 | 0 | 0 | 51.0 | | 3,820 |
| 15-17 | 0 | 0 | 49.6 | 6,220 | 49.4 | 5,118 | 100 | 4,043 | 49.0 | | 4,677 |
| Tobacco & ENDS use |  |  |  |  |  |  |  |  |  | |  |
| Never use | 92.7 | 6,118 | 81.6 | 9,580 | 79.3 | 7,202 | 76.1 | 3,069 | 84.8 | | 7,146 |
| Non-current use | 5.6 | 380 | 10.8 | 1,307 | 11.6 | 1,102 | 16.9 | 704 | 9.6 | | 839 |
| Cigarette-only use | 0.5 | 35 | 1.2 | 166 | 0.7 | 72 | 0.7 | 30 | 0.4 | | 29 |
| Non-daily | 0.4 | 33 | 1.1 | 142 | 0.6 | 59 | 0.5 | 23 | 0.3 | | 26 |
| Daily | <0.1 | 2 | 0.2 | 24 | 0.1 | 13 | 0.2 | 7 | 0.1 | | 3 |
| ENDS-only use | 1.0 | 69 | 5.2 | 604 | 7.1 | 652 | 5.4 | 204 | 4.5 | | 415 |
| Non-daily | 1.0 | 67 | 4.7 | 555 | 6.3 | 581 | 4.5 | 166 | 3.8 | | 347 |
| Daily | <0.1 | 2 | 0.4 | 49 | 0.8 | 71 | 1.0 | 38 | 0.7 | | 68 |
| Dual cigarette/ENDS user | 0.3 | 20 | 1.3 | 151 | 1.3 | 125 | 0.9 | 36 | 0.7 | | 68 |
| Non-daily cigarette, non-daily ENDS | 0.3 | 18 | 1.0 | 116 | 0.8 | 81 | 0.5 | 21 | 0.4 | | 40 |
| Non-daily cigarette, daily ENDS | <0.1 | 2 | 0.1 | 19 | 0.3 | 29 | 0.3 | 11 | 0.2 | | 25 |
| Daily cigarette, non-daily ENDS | 0 | 0 | 0.1 | 14 | 0.1 | 11 | 0.1 | 3 | <0.1 | | 2 |
| Daily cigarette, daily ENDS | 0 | 0 | <0.1 | 2 | <0.1 | 4 | <0.1 | 1 | <0.1 | | 1 |

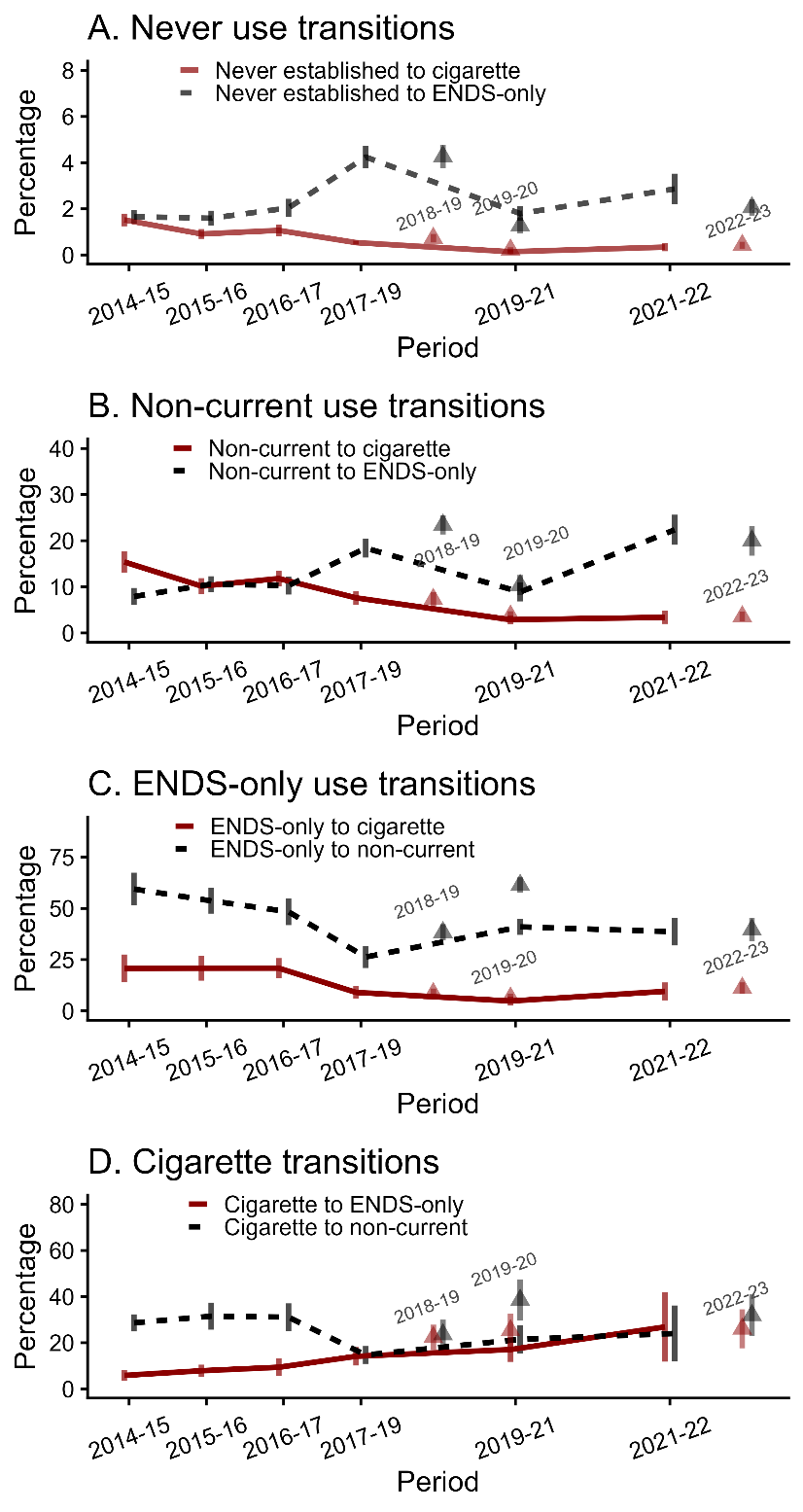

**Figure S2.** *One-year transition probabilities and 95% confidence intervals for youth with experimental cigarette and ENDS use, PATH Waves 1–7.5 (2014–22), by product use state. The x-axis represents the wave pair, and the y-axis represents the modeled probability of transition in one year. Connected points indicate standard transitions between main Waves, while triangular points correspond to transitions involving special collection Waves: 4.5–5 (2018–19), 5–5.5 (2019–20), and 7–7.5 (2022–23).*

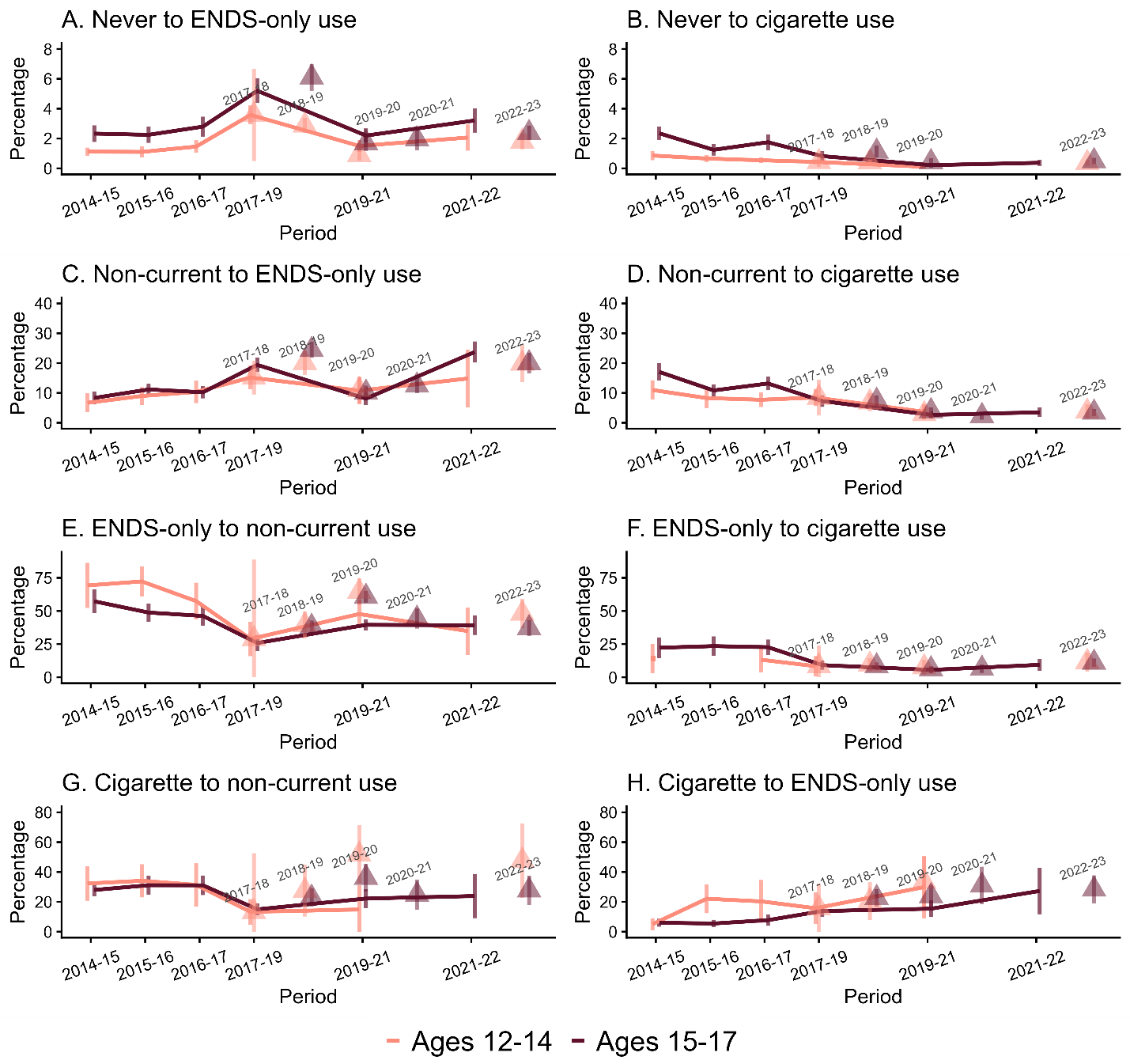

**Figure S3***. One-year transition probabilities and 95% confidence intervals for youth with experimental cigarette and ENDS use, PATH Waves 1–7.5 (2014–22), by transition and age group. The x-axis represents the year of the starting wave in each transition, and the y-axis represents the modeled probability of transition in one year. Connected and square points indicate standard transitions between main Waves, while triangular points correspond to transitions involving special collection Waves: 4-4.5 (2017-18), 4.5–5 (2018–19), 5–5.5 (2019–20), 5.5–6 (2020–21), and 7–7.5 (2022–23). Results based on fewer than five observed transitions were censored because of high uncertainty.*
